# The rise and fall of an emerging SARS-CoV-2 variant with the spike protein mutation L452R

**DOI:** 10.1101/2021.07.03.21259957

**Authors:** Orna Mor, Michal Mandelboim, Shay Fleishon, Efrat Bucris, Dana Bar-Ilan, Michal Linial, Yaniv Lustig, Israel National Consortium for SARS-CoV-2 sequencing, Ella Mendelson, Neta S. Zuckerman

**Affiliations:** Central Virology Laboratory, Public Health Services, Ministry of Health, Sheba Medical Center, Tel-Hashomer, 52621, Israel; Department of Biological Chemistry, The Alexander Silberman Institute of Life Sciences, The Hebrew University of Jerusalem, Jerusalem 91904, Israel; Department of Epidemiology and Preventive Medicine, School of Public Health, Sackler Faculty of Medicine, Tel-Aviv University, Tel-Aviv 69978, Israel

**Keywords:** SARS-CoV-2, variant, mutation, sequencing, neutralization

## Abstract

Emerging SARS-CoV-2 variants may threaten global vaccination efforts and awaited reduction in outbreak burden. In this study, we report a novel variant carrying the L452R mutation that emerged from a local B.1.362 lineage, B.1.362+L452R. The L452R mutation is associated with the Delta and Epsilon variants and was shown to cause increased infection and reduction in neutralization in pseudoviruses. Indeed, the B.1.362+L452R variant demonstrated a X4-fold reduction in neutralization capacity of sera from BNT162b2-vaccinated individuals compared to a wild-type strain. The variant infected 270 individuals in Israel between December 2020 and March 2021, until diminishing due to the gain in dominance of the Alpha variant in February 2021. This study demonstrates an independent, local emergence of a variant carrying a critical mutation, L452R, which may have the potential of becoming a variant of concern and emphasizes the importance of routine surveillance and detection of novel variants among efforts undertaken to prevent further disease spread.

## Introduction

The severe acute respiratory syndrome coronavirus 2 (SARS-CoV-2) is responsible for substantial morbidity and mortality and has been rapidly spreading worldwide since December 2019. Globally-applied Pfizer and other vaccination programs have effectively reduced the prevalence of severe disease cases and the number of hospitalizations [1][2]. However, variants of concern (VOC) such as B.1.1.7 (Alpha) [3], B.1.351 (Beta) [4], P.1 (Gamma) [5] and the most recent - B.1.617.2 (Delta) [6], have emerged in the last months. The Delta variant, first emerging in India, has been already identified in ∼100 countries worldwide. As VOC were shown to reduce vaccine efficacy in convalescent or vaccinated individuals [7][8][9], they may threaten global vaccination efforts and awaited reduction in outbreak burden. It is therefore of outmost importance to monitor such VOC and identify emerging local variants via routine sequencing from random sampling of the population.

Amongst the mutations characterizing various globally circulating variants, the L452R mutation has been recently revealed as one of the mutations associated with the most recently denoted new VOC, Delta [10]. Data analysis performed on GISAID-deposited genomes revealed that most lineages including the L452R mutation have independently emerged in recent months (since December 2020), suggesting that this mutation may significantly contribute to the adaptiveness of the virus, possibly reflecting adaptation for increasing immunity against the virus within the population [11]. Indeed, the L452R mutation is also associated with other known variants such as the A.27.1 variant identified in Mayotte [12], and the B.1.427/B.1.429 (Epsilon) variant identified in California, US [13]. The Epsilon variant became dominant in California by early 2021 and was found to be 20% more transmissible with a 2-fold increased shedding in vivo [14].

The L452 residue is located in the spike (S) protein within the receptor binding domain (RBD). Upon binding of the S protein to its human ACE-2 receptor, it forms a hydrophpbic patchon (with F490 and L492 residues) on the surface of the spike RBD which is not in direct contact with the receptor [14], [15]. Pseudoviruses with the L452R mutation were shown to have a X5.8 - 22.5 folds increase in cell entry compared to wild-type (WT) pseudoviruses [14], which was slightly lower compared to the X11.4 - 30.9 increase observed in N501Y compared to WT pseudoviruses [16]. Pseudoviruses carrying the L452R mutation were shown to be resistant to some neutralizing antibodies [17]. Accordingly, independent estimation of the Epsilon variants B.1.429 and B.1.427 carrying the L452R mutation showed a 6.7-fold and a 5.3-fold median reduction in neutralization, respectively, in comparison to the WT strain [14]. Recently, the Delta variant carrying the L452R mutation was shown to have a 5.8-fold reduction in neutralization compared to the WT strain [18].

In this study, we describe a novel emerging variant carrying the L452R mutation that emerged from a local B.1.362 lineage which was widespread in Israel prior to the entrance of the Alpha variant in December 2020. We examined the temporal and geographical distribution and the neutralization capacity of this new variant until diminishing in April 2021. Identification of this locally emerging B.1.362+L452R variant carrying a mutation known to have increased infectivity and/or reduced neutralization emphasizes the importance of routine surveillance of emerging variants, which is crucial to control further spread of SARS-CoV-2.

## Materials and Methods

### Sample collection for sequencing

Random sampling and systematic collection of PCR-positive samples for SARS-CoV-2 whole genome sequencing (WGS) was initiated in December 2020 and is now routinely conducted in Israel as part of a national effort for monitoring SARS-CoV-2 variants.

### Library preparation and sequencing

Viral genomes were extracted from 200µl respiratory samples with the MagNA PURE 96 (Roche, Germany), according to the manufacturer instructions. Libraries were prepared using COVID-seq library preparation kit, as per manufacturer’s instructions (Illumina). Library validation and mean fragment size was determined by TapeStation 4200 via DNA HS D1000 kit (Agilent). Libraries were pooled, denatured and diluted to 10pM and sequenced on NovaSeq (Illumina).

### Bioinformatics analysis

Fastq files were subjected to quality control using FastQC (www.bioinformatics.babraham.ac.uk/projects/fastqc/) and MultiQC [19] and low-quality sequences were filtered using trimmomatic [20]. Sequences were mapped to the SARS-CoV-2 reference genomes (NC_045512.2) using Burrows-Wheeler aligner (BWA) mem [21]. Resulting BAM files were sorted, indexed and subjected to quality control using SAMtools suite [22]. Coverage and depth of sequencing was calculated from sorted BAM files using a custom python script. Concensus Fasta sequences were assembled for each sample using iVar (https://andersen-lab.github.io/ivar/html/index.html), with positions with <5 nucleotides determined as Ns. Multiple alignment of sample sequences with SARS-CoV-2 reference genome (NC_045512.2) was done using MAFFT [23].

Mutation calling, translation to amino acid and identification of P681H variant sequences were done in R with a custom code using Bioconductor package Seqinr [24]. Sequences were further analyzed together with additional sequences identified as belonging to the background lineage B.1.1.50 downloaded from GISAID [25].

Phylogenetic trees were constructed using the Augur pipeline [26]. Sequences were aligned to SARS-CoV-2 reference genome (NC_045512.2) using MAFFT [23], and a time-resolved phylogenetic tree was constructed with IQ-Tree [27] and TreeTime [28] under the generalized time reversible (GTR) substitution model and visualized with auspice [26]. Lineage nomenclature was attained from Pangolin Lineages [29].

### Neutralization assays

VERO-E6 cells at concentration of 20*10^3^/well were seeded in sterile 96-wells plates with standard Eagle-Minimum Essential Medium (MEM) supplemented with 10% fetal calf serum (FCS), and stored at 37°C for 24 hours. One hundred TCID50 (median tissue culture infectious dose) of B.1.362+L452R variant, WT strain and Alpha variant isolates were incubated with inactivated sera diluted 1:10 to 1:1280 in 96 well plates for 60 minutes at 33ºC. Virus-serum mixtures were added to the VERO-E6 cells and incubated for five days at 33°C, after which Gentain violet staining (1%) was used to stain and fix the cell culture layer. Neutralizing dilution of each serum sample was determined by identifying the well with the highest serum dilution without observable cytopathic effect. A dilution equal to 1:10 or above was considered neutralizing.

## Results

### Characterization and epidemiology of the B.1.362+L452R variant

The novel B.1.362+L452R variant forms a distinct cluster in a phylogenetic tree representing randomly chosen sequenced samples from Israel (Figure 1A), constructed as part of the national program for SARS-CoV-2 whole genome sequencing and variant surveillance. The variant emerged from the B.1.362 lineage, which was one of the two most widespread lineages in Israel along with B.1.1.50 [30] until December 2020, when the Alpha variant started spreading and became the dominant variant in Israel. Although the B.1.362 lineage is detected in numerous countries, only sequences of this lineage originating in Israel formed a cluster representing the B.1.362+L452R variant. The variant is characterized by two non-synonymous mutations in the S protein, L452R (T22917G), which lies within the receptor binding domain (RBD), and L1063F (G24751T), in addition to a non-synonymous mutation in the NSP16 gene in D2179Y (G20002T) and a synonymous mutation in the M protein in G26840A. An additional characterizing mutation in the ORF3a gene - W131C (G25785T), was acquired at a later time-point and exists in most of the variant sequences (Figure 1B).

**Figure 1.**
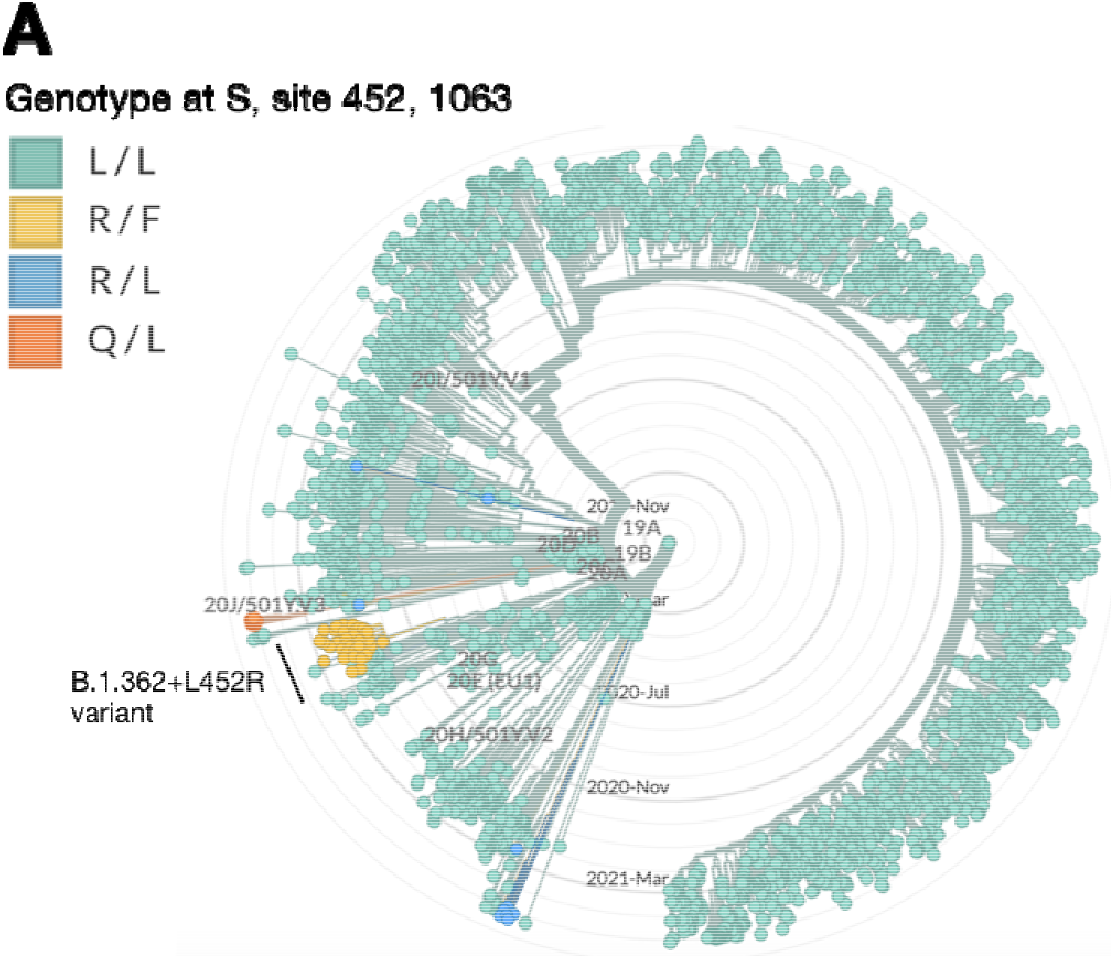
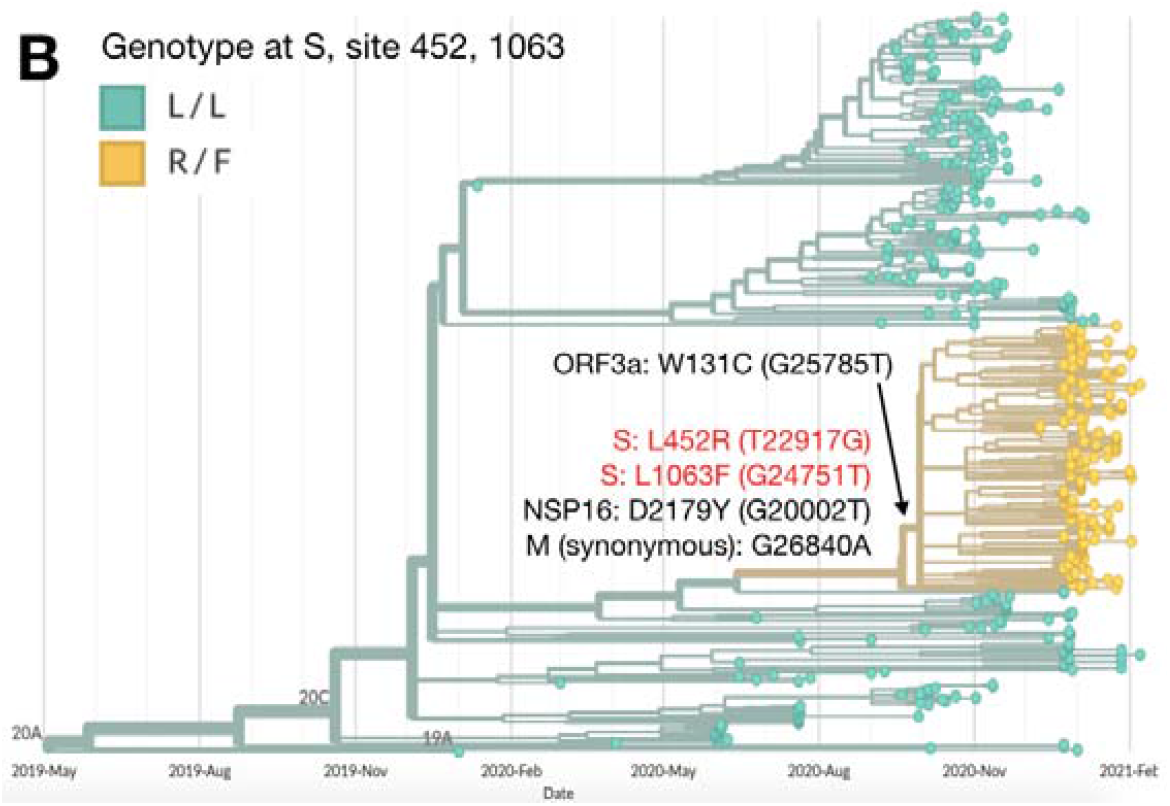
Characterization of the B.1.362+L452R variant in Israel and within the global B.1.362 lineage. (A) Phylogenetic tree representing random samples sequenced in Israel from December 2020 to May 2021 (n=3870). The genotype at the S protein in locations 452 and 1063 is indicated, with the different amino acid substitutions in these locations denoted by different colors: wild-type in green (L/L: positions 452 and 1063 are Leucine (L)) and B.1.362+L452R variant in yellow (R/F: positions 452 and 1063 were substituted to arginine (R) and phenylalanine (F), respectively). Additional variants which have a mutation in the 452 position are apparent in the tree, including a B.1.617.2 variant cluster with R/L in blue and a C.37 variant cluster with Q/L in orange. (B) Global SARS-CoV-2 genomes from of the B.1.362 lineage only (n=441). The B.1.362+L452R variant cluster is composed of Israeli viruses only. Phylogenetic trees were created with Nextstrain Augur pipeline and visualized with Auspice [26]. Non-Israeli B.1.362 lineage sequences were downloaded from GISAID [25] and identified with Pangolin [6].

Until April 2021, an overall of 270 individuals were infected with the B.1.362+L452R variant. Most infected individuals were randomly sampled from various districts across Israel, with no age or gender bias. The prevalence of the variant in the population gradually decreased with time, from 6% out of 854 randomly sequenced sampled in December 2020, 0% out of 1604 random samples in April 2021 (Table 1). This decrease was also observed in the B.1.362 lineage that decreased from 19% in December 2020 to 0% in April 2021, due to the penetration of the Alpha variant in December 2020 which became the dominant variant in Israel since February 2021 (Figure 2).

**Table 1.** Epidemiology of the B.1.362+L452R variant. Frequency and patient-related information of the B.1.362+L452R variant. Ret. abroad is return from abroad; follow vacc is following vaccination; Jud & Sam is Judea and Samaria; hospital stat is hospital status.

|  |  | December<br>2020 | January<br>2021 | February<br>2021 | March<br>2021 | April<br>2021 |
| --- | --- | --- | --- | --- | --- | --- |
| gender | Male | 10 | 37 | 54 | 3 |  |
|  | Female | 17 | 63 | 40 | 1 |  |
|  | Unknown | 24 | 20 | 1 |  |  |
| age |  | 35±25 | 30±18 | 30±21 | 59±29 |  |
| reason | random | 49 | 118 | 95 | 3 |  |
|  | ret. abroad | 2 | 2 |  |  |  |
|  | follow vacc |  |  |  | 1 |  |
| district | Northern | 2 | 8 |  |  |  |
|  | Haifa |  | 2 | 5 | 1 |  |
|  | Central | 6 | 32 |  |  |  |
|  | Tel Aviv | 10 | 20 |  |  |  |
|  | Jerusalem | 7 | 37 |  | 1 |  |
|  | Jud & Sam |  | 1 | 1 |  |  |
|  | Southern | 1 | 6 | 26 |  |  |
|  | Unknown | 25 | 14 |  | 2 |  |
| hospital stat | not hospitalized | 50 | 120 | 93 | 4 |  |
|  | not critical |  |  |  |  |  |
|  | critical | 1 |  | 1 |  |  |
|  | deceased |  |  | 1 |  |  |
| total B.1.362+L452R |  | 51 | 120 | 95 | 4 | 0 |
| % of random |  | 5.97 | 4.76 | 3.83 | 0.24 | 0.00 |

**Figure 2.**
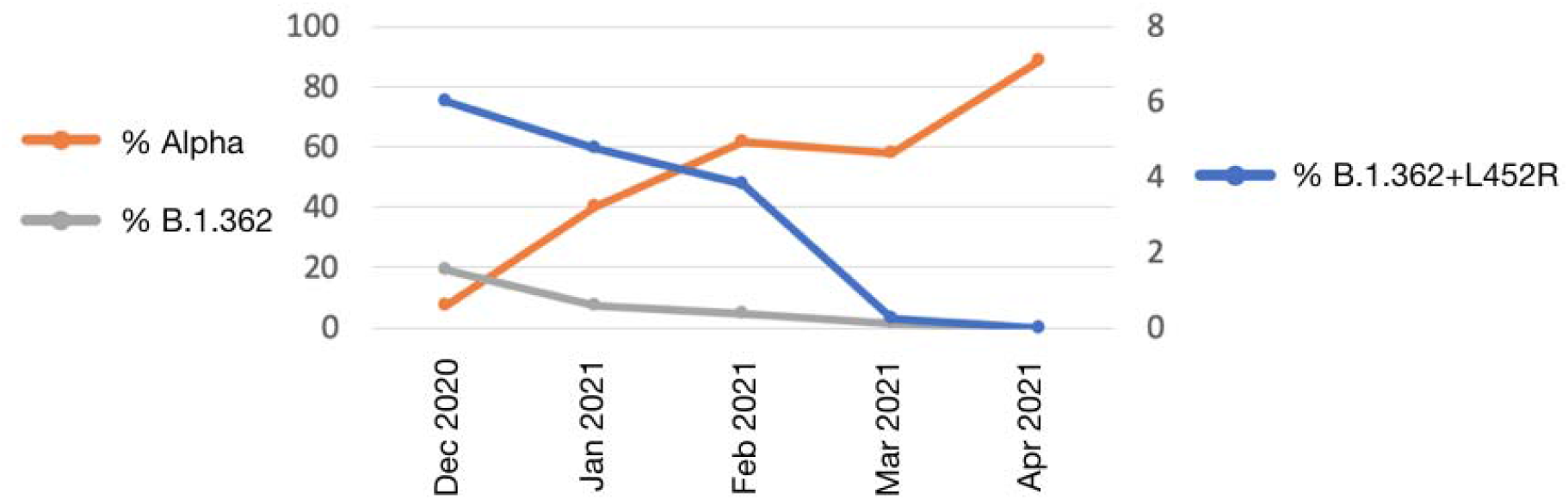
Prevalence of the B.1.362+L452R variant and other lineages in Israel. The prevalence of the B.1.362+L452R variant is denoted in blue (right y-axis); the Alpha variant is denoted in orange (left y-axis); the B.1.362 lineage is denoted in grey (left y-axis).

### Neutralizing capacity against the B.1.362+L452R variant following BNT162b2 vaccination

The neutralization potency of sera from vaccinated individuals against the B.1.362+L452R variant was compared to the neutralization capacity against two other strains - the Alpha variant (isolate hCoV-19/Israel/CVL-46879-ngs/2020) and a local WT strain from Israel (isolate hCoV-19/Israel/CVL-45526-ngs/2020). VERO-E6 cells were infected with each viral strain and pre-incubated with serial dilutions of serum samples obtained from individuals >30 days after having received the second dose of the Pfizer BNT162b2 vaccine. Results show that the B.1.362+L452R variant had X2.3 and X4 folds reduced neutralization capacity compared to the Alpha variant and the Israeli WT strain, respectively (Figure 3).

**Figure 3.**
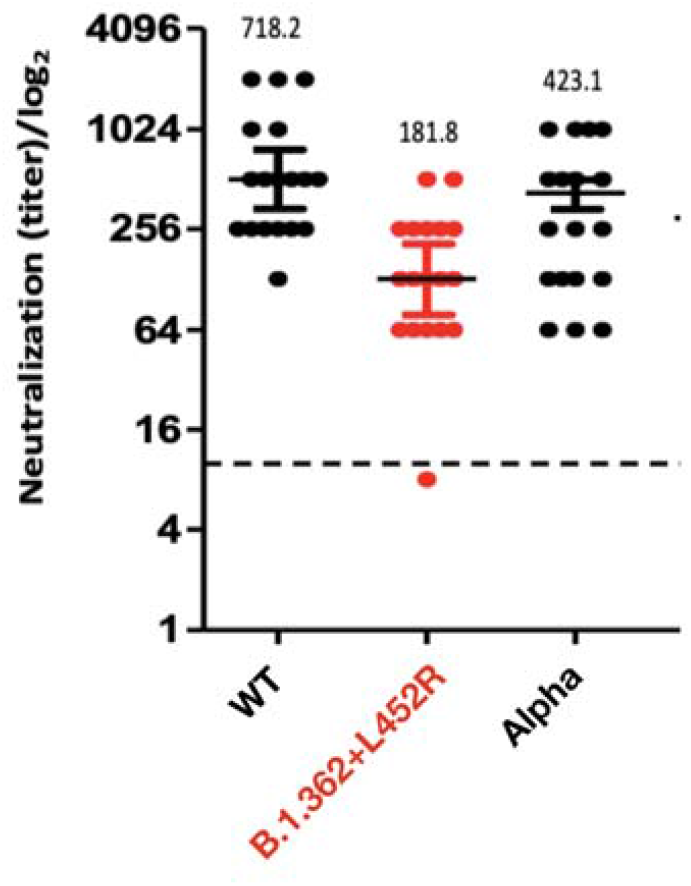
Neutralization capacity of the B.1.362+L452R variant. Neutralization assays were carried out with VERO-E6 cells infected with the B.1.362+L452R variant, the Alpha variant and a local WT strain, using sera from individuals obtained at least 30 days following the second dose of the Pfizer-BionTech vaccine. Bars represent the log geometric mean titer (GMT) and 95% confidence intervals. A dilution equal to 1:10 or above was considered neutralizing (dashed line).

## Discussion and conclusions

The S protein mutation L452R has been recently in the spotlight as one of the main mutations within the B.1.617 lineage variants originating in India, most notably the Delta variant which is currently uninhibitedly spreading worldwide and is suspected to be one of the most contagious SARS-CoV-2 variants yet [10]. It is estimated to be 60% more transmissible than the Alpha variant, where the high transmissibility is attributed to the combination of three main mutations in the S protein: E484Q, L452R and P681R [31]. Herein, we report a novel variant carrying the S protein L452R mutation, which emerged from a local B.1.362 lineage. The variant was identified in 270 individuals overall, however its prevalence, which was 6% of randomly sequenced samples in December 2020, gradually declined and in April 2020 it was not observed in any of the 1604 samples sequenced that month (Figure 2). The B.1.362 lineage from which the B.1.362+L452R variant emerged, was one of the two most prevalent SARS-CoV-2 lineages (along with B.1.1.50) in Israel since the start of the epidemic in Israel in March 2020. However, with the penetration of the dominant Alpha variant in December 2020, all local lineages started to decline, including the B.1.362 lineage and the B.1.362+L452R variant. In addition, as a result of the successful vaccination program in Israel that started already in December 2020, the number of overall infected individuals drastically declined, such that by April 2021 the B.1.362+L452R variant was completely diminished.

The identification of this locally-emerging variant carrying the L452R mutation, accompanied by additional independently-emerging variants carrying the same mutation (Delta and additional B.1.617 lineage variants, Epsilon, A.27.1), emphasizes the relevance of the L452R mutation to the virus adaptiveness. In addition, viral variants carrying the L452R mutation such as the Epsilon and Delta variants have been shown to have a 6.7 and a 5.8-fold reduction in neutralization against WT virus, respectively [14][18]. Similarly, this study demonstrated a X4-fold decrease in neutralization capacity of sera derived from fully vaccinated individuals against the B.1.362+L452R variant, suggesting that it is this mutation that accounts for the impact on the neutralization potential of the different variants that carry the L452R mutation.

The B.1.362+L452R variant is currently not defined as a variant of concern in Israel, despite the observed reduction in neutralization, as it is no longer observed via routine sequencing since April 2021. Nevertheless, this study emphasizes the importance of SARS-CoV-2 continuous surveillance which proved to be essential for the identification and monitoring of emerging variants even in countries that reached a high level of vaccination. Specifically, emerging variants carrying mutations that are known in VOC associated with increased infectivity and/or reduction in neutralization, such as L452R, and the accompanying concern that such variants will gain additional mutations facilitating enhanced and rapid spread – locally and worldwide.

## Data Availability

All sequence data has been deposited and available in GISAID

## Acknowledgments

The National Covid-19 Information & Knowledge Center. The Department of Laboratories, Public Health Services, Ministry of Health.

## Notes

### Competing Interest Statement

The authors have declared no competing interest.

### Funding Statement

No external funding was received

### Author Declarations

The study was conducted according to the guidelines of the Declaration of Helsinki, and approved by the Institutional Review Board of the Sheba Medical Center institutional review board (7045-20-SMC). Patient consent was waived because the study used remains of clinical samples and the analysis used anonymous clinical data.

